# Causal Analysis of Health Interventions and Environments for Influencing the Spread of COVID-19 in the United States of America

**DOI:** 10.1101/2020.09.29.20203505

**Authors:** Zhouxuan Li, Tao Xu, Kai Zhang, Hong-Wen Deng, Eric Boerwinkle, Momiao Xiong

## Abstract

As of August 27, 2020, the number of cumulative cases of COVID-19 in the US exceeded 5,863,363 and included 180,595 deaths, thus causing a serious public health crisis. Curbing the spread of Covid-19 is still urgently needed. Given the lack of potential vaccines and effective medications, non-pharmaceutical interventions are the major option to curtail the spread of COVID-19. An accurate estimate of the potential impact of different non-pharmaceutical measures on containing, and identify risk factors influencing the spread of COVID-19 is crucial for planning the most effective interventions to curb the spread of COVID-19 and to reduce the deaths. Additive model-based bivariate causal discovery for scalar factors and multivariate Granger causality tests for time series factors are applied to the surveillance data of lab-confirmed Covid-19 cases in the US, University of Maryland Data (UMD) data, and Google mobility data from March 5, 2020 to August 25, 2020 in order to evaluate the contributions of social-biological factors, economics, the Google mobility indexes, and the rate of the virus test to the number of the new cases and number of deaths from COVID-19. We found that active cases/1000 people, workplaces, tests done/1000 people, imported COVID-19 cases, unemployment rate and unemployment claims/1000 people, mobility trends for places of residence (residential), retail and test capacity were the most significant risk factor for the new cases of COVID-19 in 23, 7, 6, 5, 4, 2, 1 and 1 states, respectively, and that active cases/1000 people, workplaces, residential, unemployment rate, imported COVID cases, unemployment claims/1000 people, transit stations, mobility trends (transit), tests done/1000 people, grocery, testing capacity, retail, percentage of change in consumption, percentage of working from home were the most significant risk factor for the deaths of COVID-19 in 17, 10, 4, 4, 3, 2, 2, 2, 1, 1, 1, 1 states, respectively. We observed that no metrics showed significant evidence in mitigating the COVID-19 epidemic in FL and only a few metrics showed evidence in reducing the number of new cases of COVID-19 in AZ, NY and TX. Our results showed that the majority of non-pharmaceutical interventions had a large effect on slowing the transmission and reducing deaths, and that health interventions were still needed to contain COVID-19.

## Introduction

As of August 27, 2020, the number of cumulative cases of COVID-19 in the US exceeded 5,863,363 and included 180,595 deaths, thus causing a devastating public health and economic crisis. Since the number of new cases in the US remains high (43,814 in the US on August 27, 2020), curbing the spread of COVID-19 is urgently needed (Callaway 2020). There is increasing recognition that many geographic, economic and environmental factors contribute to the outbreak of COVID-19. In the absence of vaccines and specifically effective medications, non-pharmaceutical public health interventions and personal hygiene practices are the only options to slow the spread of COVID-19 (Priyadarsini and Suresh, 2020; Irfan 2020). The effects of the different factors and intervention measures on the spread of COVID-19 vary. Identifying key factors that most contribute to the rapid spread of COVID-19, and accurately estimating the potential impact of different non-pharmaceutical measures for containing COVID-19 are crucial for planning the most effective interventions to curb the spread of Covid-19 (Farseev et al. 2020).

The widely used statistical methods for COVID-19 epidemiological factor analysis and evaluation of intervention measures include correlation analysis (Farseev et al. 2020; Tantrakarnapa et al. 2020; Priyadarsini and Suresh, 2020; Nakada and Urban 2020), regression (Chaudhry et al. 2020), the spatial autoregressive (SAR) model (Baum and Henry, 2020), logistic regression (Coccia M 2020) and a transmission dynamic model coupled with a linear model (Livadiotis 2020). The most examined scalar factors consists of underlying health conditions such as high blood pressure, diabetes, stroke, cardiac or kidney diseases, and aging individuals (Priyadarsini and Suresh, 2020; Zhou et al. 2020; Raghupathi 2019), atmospheric temperature (Tantrakarnapa et al. 2020), age, gender, ethnicity, and population density (Priyadarsini and Suresh, 2020; Anderson et al. 2020), airflow (Priyadarsini and Suresh, 2020), and socioeconomics such as median income (Coccia 2020; Saadat et al. 2020).

The most explored non-pharmaceutical public health interventions and digital technologies for curbing the spread of COVID-19 include social distancing, case isolation and quarantine as well as closuring borders, schools travel restrictions, use of face-masks, and testing (Flaxman et al. 2020; Viner et al. 2020; Quilty et al. 2020; Ngonghala et al. 2020) andpopulation surveillance, case identification, contact tracing, mobility data collection, and communication technology, which utilize billions of mobile phones and large online datasets to provide information for the evaluation of intervention strategies and to strengthen the curb of the spread of COVID-19 (Budd et al. 2020; Ngonghala et al. 2020; Badawy and Radovic 2020).

Although association analysis is of great importance for curbing the spread of COVID-19, association measures dependence between two variables or two sets of variables in the data, and use the dependence for prediction and evaluation of the effects of environmental, social-economic factors and public health interventions on the spread of COVID-19 (Altman and Krzywinski 2015; Sharkey and Wood 2020). It is well recognized that association analysis is not a direct method to discover the causal mechanism of complex diseases. Association analysis may detect superficial patterns between intervention measures and transmission variables of COVID-Its signals provide limited information on the causal mechanism of the transmission dynamics of COVID-19 (Steigera et al. 2020). Association analysis has been a major paradigm for statistical evaluation of the effects of influencing factors and health interventions on the spread of COVID-19 (Li et al. 2020). Understanding the transmission mechanism of COVID-19 based on association analysis remains elusive. The question to uncover the transmission mechanisms of COVID-19 is causal in nature.

Distinguishing causation from association is an age-old problem. Methods for causation analysis that is one of the most challenging problems in science and technology need to be developed as an alternative to association analysis (Zenil et al., 2019).A number of researchers have performed causal analysis of COVID-19 to evaluate the causal effects of mobility, awareness, and temperature (Steigera et al. 2020), social distancing (Sharkey and Wood 2020), mobility (Ramachandra and Sun 2020), herd immunity (Friston et al. 2020), and mask use (Chernozhukov et al. 2020). However, most causal analysis of COVID-19 have treated time series data as pseudo-cross sectional data. In some cases causal analysis of COVID-19 treated the data as time series; time series was assumed stationary. In practice, the number of new cases and the number of deaths from COVID-19 were nonstationary time series in most cases. The environmental, social-economic and geographic factors, and intervention measures include two types of data: scalar variables and time series (stationary or nonstationary) variables.

The purpose of this paper is to develop a general framework for the causal analysis of COVID-19 in the US. The number of new cases and deaths from COVID-19 are taken as response variables. The factors and intervention measures are taken as potential causal variables. If the factor and intervention variables are scalar variables, the additive noise models (AMMs) (Peters et al. 2014) are used to test for causation between the response variable and potential causal variable where the number of new cases or deaths should be averaged over time. Most intervention measures are time series data. An essential difference between time series and cross-sectional data is that the time series data have temporal order, but cross sectional data do not have any order. As a consequence, the causal inference methods for cross sectional data cannot be directly applied to time series data. Basic tools in statistical analysis are the raw of large numbers and the central limit theorem. Applications of these tools usually assume that all moment functions are constant. When the moment functions of the time series vary over time, the raw of large numbers and the central limit theorem cannot be applied. In order to use basic probabilistic and statistical theories, the nonstationary time series must be transformed to stationary time series (Johansen 1991).

A widely used concept of causality for time series data is Granger causality (Granger 1969; Eichler 2013). Underlying the Granger causality is the following two principles:

1. Effect does not precede the cause in time;
2. The effect series contains unique causal series information which is not present elsewhere.

The multivariate linear Granger causality test will be used to test causality between the number of new cases and deaths from COVID-19 and environmental, economic and intervention time series variables (Bai et al. 2010). The proposed ANMs and multivariate linear Granger causality analysis methods are applied to the surveillance data of lab-confirmed Covid-19 cases in the US, UMD data, and Google mobility data from March 5, 2020 to August 25, 2020 in order to evaluate the contributions of social-biological factors, economics, the Google mobility indexes, and the rate of virus testing to the number of the new cases and number of deaths from COVID-19.

## Materials and Methods

### Nonlinear additive noise models for bivariate causal discovery

The ANMs are used for identifying causal effect of a factor or an intervention measure on the number of new cases or an intervention measure (Peters et al. 2014; Jiao et al. 2018). Assume no confounding, no selection bias, and no feedback. Let be the average number of new cases or new deaths from COVID-19 and be a scalar factor or an intervention measure such as gender, population density, ethnic group, among others. Consider a bivariate additive noise model *X* → *Y* where *Y* is a nonlinear function of *X* and independent additive noise *E*_*Y*_ :

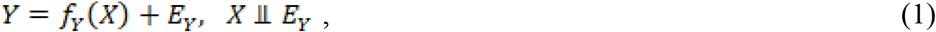

where *X* and *E*_*Y*_ are independent. Then, the density *p*_*X,Y*_ is said to be induced by the additive noise models (ANM) from *X* to *Y* (Mooij et al. 2016). In some cases, we may have the following alternative direction of the ANMs: *Y* → *X* :

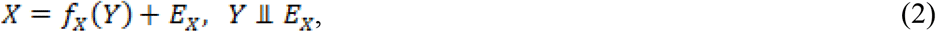

where *Y* and *E*_*X*_ are independent. If the density *P*_*X,Y*_ is induced by the ANM *X* → *Y*, but not by the ANM *X* → *Y*, then the ANM *X* → *Y* is identifiable.

Assume that *n* +*m* state data were sampled. Divide the dataset into a training data set by specifying *D*_1_ = {*Y_n_, X_n_*},*Y_n_* = [*y*_1_, …,*y_n_*]*^T^, X_n_* = [*x*_1_, …,*x_n_*]*^T^* for fitting the model and a test data set 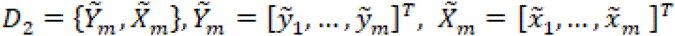 for testing the independence, where *n* is not necessarily equal to *m*.

Procedures for using the ANM to assess causal relationships between two variables are summarized below (Jiao et al. 2018).

**Step 1**. Regress *Y* on *X* using the training dataset *D*_1_ and non-parametric regression methods:

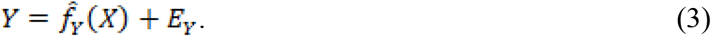

**Step 2**. Calculate the residual 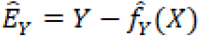 using the test dataset *D*_2_ and test whether the residual 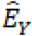 is independent of causal *X* to assess the ANM *X* → *Y*.

**Step 3**. Repeat the procedure to assess the ANM *Y*→ *X*.

**Step 4**. If the ANM in one direction is accepted and the ANM in the other is rejected, then the former is inferred as the causal direction.

There are many non-parametric methods that can be used to regress *Y* on *X* or regress *X* on *Y*. For example, we can use neural networks (Heydari et al. 2019), smoothing spline regression methods (Wang 2011), B-spline (Wang 2017) and local polynomial regression (LOESS, see Cleveland, 1979). In this paper, the smoothing spline regression method was used to fit the regression models.

Covariance can be used to measure association, but cannot be used to test independence between two variables with a non-Gaussian distribution. A covariance operator that is a generalization of the finite dimensional covariance matrix to infinite dimensional feature space can be used to test for independence between two variables with arbitrary distributions. Specifically, we will use the Hilbert-Schmidt norm of the cross-covariance operator or its approximation, the Hilbert-Schmidt independence criterion (HSIC) to measure the degree of dependence between the residuals and potential causal variable and test for their independence (Gretton et al. 2005; Mooij et al. 2016).

The covariance operator can be defined as

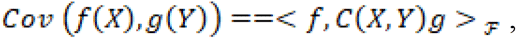

where *f, g* any nonlinear functions and *C*(*X, Y*) is the covariance operator and <.> _ℱ_ is an inner product in the ℱ Hilbert space. The Hilbert-Schmidt norm of the covariance operator can be used as criterion for assessing independence between two random variables and is called the Hilbert-Schmidt independence criterion (HSIC). The Hilbert-Schmidt norm of the centered covariance operator is defined as

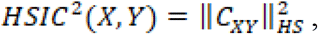

where ‖ · ‖ _*HS*_ is the Hilbert-Schmidt norm.

We know that (Wang et al. 2018)

*HSIC*^2^(*X,Y*) =0 if and only if *X* and are *Y* independent.

*HSIC*^2^(*X,Y*) can be approximated by

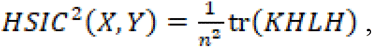

where *n* is a sample size, *K* and *L* are *n* × *n* dimensional kernel matrices and 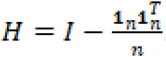. We used the Gaussian kernel: 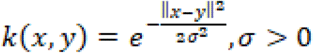 to test independence between the potential cause *X* and residual *E*_*Y*_; we calculated *HSIC*^2^(*X, E_Y_*) as follows.

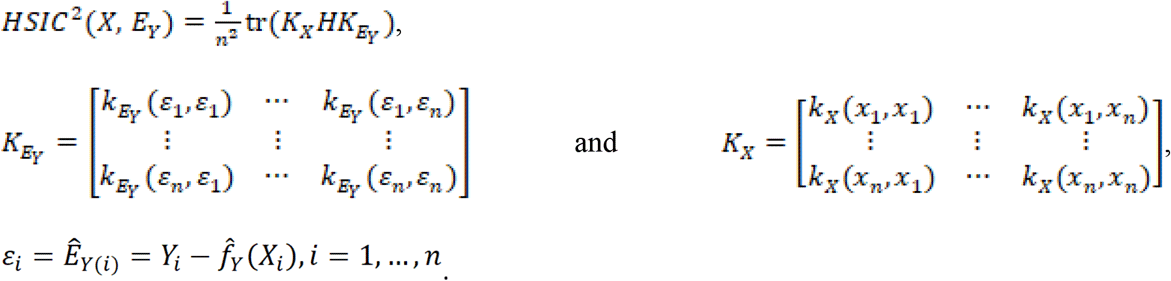

In summary, the general procedure for testing independence between the average number of new cases or new deaths and the scalar factor or intervention measure is given as follows (Mooij et al. 2016; Jiao et al. 2018):

**Step 1**: Divide a data set into a training data set *D*_1_ = {*Y*_*n*_,*X*_*n*_}for fitting the model and a test data set 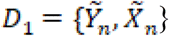 for testing the independence.

**Step 2:** Use the training data set and smoothing spline regression nonparametric regression methods

a. Regress *Y* on *X* :*Y*= *f* _*Y*_(*x*)+*E*_*Y*_,
b. Regress *X* on *Y* : *X* = *f* _*X*_ (*y*) + *E*_*X*_.

**Step 3:** Use the test data set and estimated smoothing spline regression nonparametric regression that fits the test data set *D*_2_={*Y*_*n*_, *X*_*n*_} to predict residuals:

a. 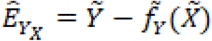,
b. 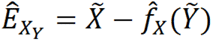.

**Step 4:** Calculate the dependence measures *HSIC*^2^ (*E* _*Y*_, *X*) and *HSIC*^2^ (*E* _*X*_,*Y*).

**Step 5:** Infer causal direction:

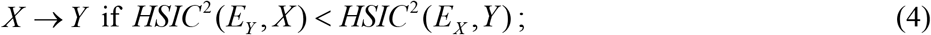

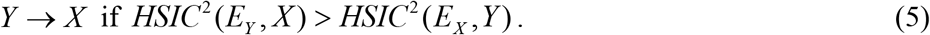

If *HSIC* ^2^ (*E*_*Y*_, *X*) = *HSIC* ^2^ (*E*_*X*_, *Y*), then causal direction is undecided.

We do not have closed the analytical forms for the asymptotic null distribution of the HSIC and hence it is difficult to calculate the P-values of the independence tests. To solve this problem, the permutation/bootstrap approach can be used to calculate the P-values of the causal test statistics. The null hypothesis is

*H*_*0*_: no causations *X* → *Y* and *Y* → *X* (Both *X* and *E*_*Y*_ are dependent, and *Y* and *E*_*X*_ are dependent).

Calculate the test statistic:

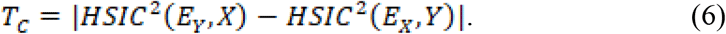

Assume that the total number of permutations is *np*. For each permutation, we fix *X*_*i*_,*i*=1, …, *n* and randomly permutate *Y*_*i*_,*i* = 1 …, *n* Then, fit the ANMs and calculate the residuals 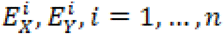 and test statistic *T*_*c*_. Repeat above procedures *n*_*p*_ times. The P-values are defined as the proportions of the statistic 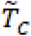 (computed on the permuted data) greater than or equal to 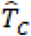 (computed on the original test data *D*_*2*_).

### Multivariate Linear Granger Causality Test

Before performing multivariate linear Grander causality test, we first need to transform nonstationary time series to stationary time series.

Consider an *m*-variable VAR with *p* lags:

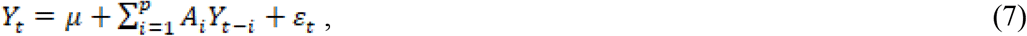

Where *Y*_*t*_ is a dimensional vector, the *A*_*i*_ (*I* = 1,.., *p*) are *m* × *m* coefficient matrices and m dimensional residual vector *ε*_*t*_, is assumed to have mean zero (*E*[*ε*_*t*_ = 0, with no autocorrelation 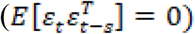, but can be correlated across equations 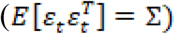.

Vector error correction model (VECM) consists of first differences of cointegrated *I* (1) variables, their lags, and error correction terms:

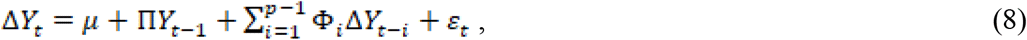

where matrixes П and Ф_*i*_(*i*=1,…,*p*-1) are functions of matrices *A*_*i*_(*i*=1,…,*p*).

When two non-stationary variables are cointegrated, the VAR model should be augmented with an error correction term for testing the Granger causality (Engle and Granger, 1987).

The VECM can be reduced to

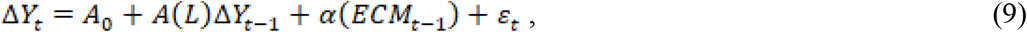

Where

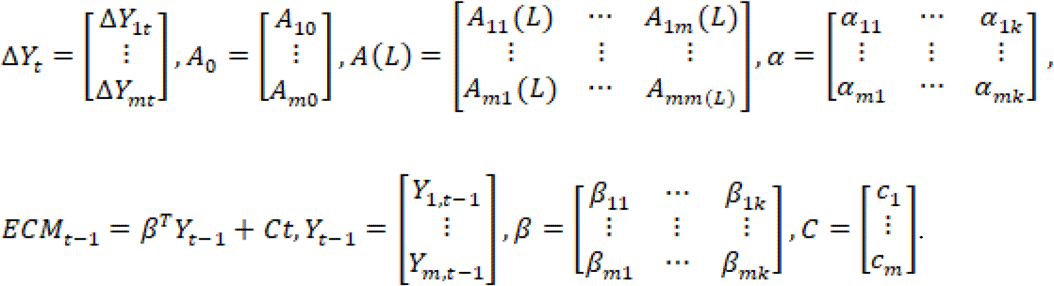

Consider two non-stationary time series, *X*_*t*_=[*X*_1*t*_,*…,X* _*m,t*_]^*T*^ and *Y*_*t*_=[*Y*_1*t*_, *…,Y* _*mt*_]^*T*^. Let *m*=*m*_1_ + *m*_2_.

Suppose that *X*_*t*_ and *Y*_*t*_ are cointegrated with the residuals *ECM*_*t*_. The VECM model for testing the Granger causality is given by

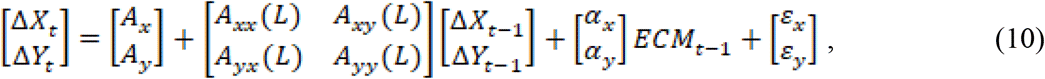

where *A*_*x*_ and *A*_*y*_are *m*_1_ and *m*_2_ dimensional vectors of intercept terms, respectively, *A*_*xx*_(*L*), *A*_*xy*_(*L*), *A*_*yx*_(*L*) and *A*_*yy*_(*L*) are *n*_1_ × *n*_1_, *n*_1_ × *n*_2_, *n*_2_ × *n*_1_ and *n*_2_ × *n*_2_ dimensional matrices of lag polynomials, respectively, and α_*x*_ and α_*y*_ are *n*_1_ and *n*_2_ dimensional coefficient vectors for the error correction term *ECM*_*t*-1_, respectively. The lag length was selected using the two-stage procedure (Abdalla and Murinde, 1997).

There are four different cases of causal relationships between two vectors of time series *X*_*t*_ and *Y*_*t*_ (Bai et al. 2010)

1. If *A*_*xy*_ (*L*) is significantly different from zero, while *A*_*yx*_ (*L*) shows no significantly different from zero, then there exists a unidirectional Ganger causality from time series *Y*_*t*_ to *X*_*t*_;
2. If *A*_*yx*_ (*L)* is significantly different from zero, while *A*_*xy*_ (*L*) shows no significantly difference from zero, then there exists a unidirectional Ganger causality from *X*_*t*_ to *Y*_*t*_;
3. If both coefficients *A*_*xy*_ (*L*) and *A*_*yx*_ (*L)* are significantly different from zero, then there exists bidirectional Granger causality between *X*_*t*_ and *Y*_*t*_;
4. If both coefficients *A*_*xy*_ (*L*) and *A*_*yx*_ (*L)* are not significantly different from zero, then *X*_*t*_ and *Y*_*t*_ are not rejected to be independent.

The four statements imply that Ganger causal relationships between *X*_*t*_ and *Y*_*t*_ depend on the coefficients *A*_*xy*_ (*L*) and *A*_*yx*_ (*L)*. Therefore, the null hypotheses for testing the Ganger causality between *X*_*t*_ and *Y*_*t*_ are

1. 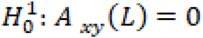,
2. 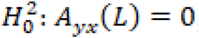, and
3. Both 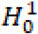 and 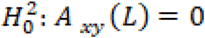 and *A*_*xy*_ (*L*) = 0.

Likelihood ratio tests for multivariate Granger causality are given by the following.

1. The likelihood ration statistics for testing the null hypothesis: 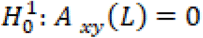 is

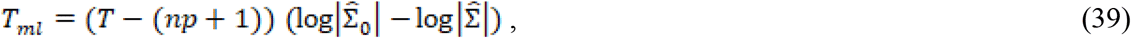

which is asymptotically distributed as a central 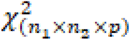 under the null hypothesis 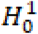.
2. The likelihood ration statistics for testing the null hypothesis: 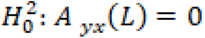 is

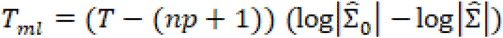

which is asymptotically distributed as a central 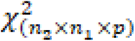 under the null hypothesis 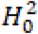.
3. The likelihood ration statistics for testing the null hypothesis: 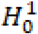 and 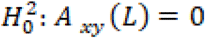 and *A*_*yx*_ (*L)=*0 is

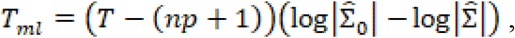

which is asymptotically distributed as a central 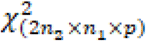 under the null hypothesis 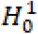 and 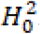.

## Data Collection

Data on the number of new cases and new deaths of Covid-19 across the 50 states in the US were obtained from John Hopkins Coronavirus Resource Center (https://coronavirus.jhu.edu/MAP.HTML). Google mobility indexes were downloaded from Google COVID-19 Community Mobility Reports (https://www.google.com/covid19/mobility/). Comprehensive data and insights on COVID-19’s impact on mobility, economy, and society were downloaded from the University of Maryland COVID-19 Impact Analysis Platform (https://data.covid.umd.edu) (Maryland Transportation Institute, 2020; Zhang et al. 2020). All data were collected from March 5, 2020 to August 25, 2020.

## Results

### Test for scalar potential causes

The scalar variables tested for causation of the new cases and deaths from COVID-19 in the US included the number of contact tracing workers per 100,000 people, percent of population above 60 years of age, median income, population density, percentage of African Americans, percentage of Hispanic Americans, percentage of males, employment density, number of points of interests for crowd gathering per 1000 people, number of staffed hospital beds per 1000 people, and number of ICU beds per 1000 people. The number of new cases and deaths were averaged over time. Each state was a sample. Since the sample sizes were small, the P-value for declaring significance was 0.05 without Bonferroni correction for multiple comparison. The P-values for testing 11 scalar potential causes of the number of new cases and deaths from COVID-19 in the US were summarized in Table 1. We observed from Table 1 that population density (P-value < 0.0002) and percentage of males (P-value < 0.03) showed significant evidence of causing the spread of COVID-19. Percentage of Hispanic Americans (P-value < 0.0575) was close to significance. Percentage of African American (P-value < 0.024) and population density (P-value < 0.025) showed significant evidence of causing deaths due to COVID-19. P-values of employment density (P-value < 0.059) and percentage of Hispanic Americans (P-value < 0.064) were close to significance level 0.05 for causing death.

**Table 1.** P-values for testing 11 scalar potential causes of the number of new cases and deaths from COVID-19 in the US.

| Risk Factor | P-value |  |
| --- | --- | --- |
|  | New cases | Deaths |
| Percent of population above the age of 60 years of age | 0.4634 | 0.1530 |
| Median income | 0.1109 | 0.0760 |
| Percentage of African Americans | 0.6526 | 0.0240 |
| Percentage of Hispanic Americans | 0.0575 | 0.0640 |
| Percentage of males | 0.0300 | 0.1440 |
| Population density | 0.0002 | 0.0250 |
| Employment density | 0.4571 | 0.0590 |
| Number of points of interests for crowd gathering per 1000 people | 0.3224 | 0.5810 |
| Number of staffed hospital beds per 1000 people | 0.6732 | 0.3130 |
| Number of ICU beds per 1000 people, | 0.5134 | 0.4860 |
| Number of contact tracing workers per 100,000 people | 0.4203 | 0.8190 |

The second most significant demographic risk factor for the spread of COVID-19 was percentage of males. We found higher COVID-19 morbidity in males than females. However, we did not find higher COVID-19 mortality in males than females.

Population density was an important risk factor for both the spread and death from COVID-High density resulted in closer contact, stronger interaction among residents and lower social distancing, which facilitated the spread and increased the death rate from COVID-19 (Rocklöv and Sjödin 2020; Pequeno et al. 2020; Henderson 2020; Rajan et al. 2020). However, our results were contradictory with the conclusion of Hamidi et al. (2020). Some literature also confirmed that high proportion of African Americans caused a high rate of deaths (Rajan et al. 2020; Golestaneh et al. 2020; Mahajan and Larkins-Pettigrew 2020). Our results concluded that percentage of Hispanic Americans was a weak risk factor for both the spread and death fromCOVID-19, while the literature showed stronger evidence that Hispanic communities were highly vulnerable to COVID-19 (Calo et. Al. 2020).

It was reported that higher COVID-19 mortality in males than females can be due to the following factors (Bwire 2020). The first factor was higher expression of angiotensin-converting enzyme-2 (ACE 2; receptors for coronavirus) in males than females. The second factor was sex-based immunological differences due to sex hormone and the X chromosome.

### Test for Granger Causality

Daily mobility and social distancing data from a COVID-19 impacted the analysis platform, including four categories: category A: mobility and social distancing, category B: COVID and health, category C: economic impact, and category D: vulnerable population. A total of 12 temporal metrics in four categories and 12 metrics from the COVID-19 impact analysis platform, six daily Google Community Mobility indexes and protest attendee data that captured real-time trends in movement patterns for each state in the US were included in the analysis to test for Granger causality between these risk factors, health intervention measures and the number of new cases and deaths from COVID-19 across 50 states in the US (Zhang et al. 2020; Google community mobility reports, 2020). The total number of variables to be tested was 19. The P-value for declaring significance after Bonferroni correction was 0.0025.

All 19 metrics except for protest attendee showed high significance in causing a reduction of the new cases of COVID-19 in 19 less affected states: VT, WY, ME, AK, NH, WV, ND, SD, NM, RI, DE, KY, KS, CT, CO, IA, WA, WI, and MS. Most of these states were less populated. However, although CA was most affected and the most populated state, all 19 metrics except protest attendance showed a strong significance in causing rapid spread of COVID-19 (Table 2 and Table S1).

**Table 2.** P-values testing 18 temporal potential causes of the number of new cases of COVID-19 in the top 10 most affected states and bottom 10 less affected states in the US.

| State | CA | FL | TX | NY | GA | IL | AZ | NJ | NC | TN |
| --- | --- | --- | --- | --- | --- | --- | --- | --- | --- | --- |
| Number of cumulative cases | 673095 | 605502 | 586730 | 430774 | 258354 | 224887 | 199273 | 190021 | 157741 | 145417 |
| Miles/person | 7.7E-08 | 3.1E-02 | 2.3E-03 | 9.1E-02 | 2.8E-04 | 4.9E-04 | 7.3E-05 | 4.7E-03 | 9.2E-06 | 5.9E-06 |
| Population | 1.6E-06 | 3.6E-02 | 5.1E-03 | 6.1E-02 | 3.2E-04 | 5.2E-05 | 1.9E-04 | 1.2E-03 | 2.5E-04 | 2.9E-05 |
| % change in consumption | 1.4E-05 | 5.5E-02 | 4.7E-03 | 1.8E-03 | 8.7E-04 | 1.8E-04 | 2.2E-04 | 9.6E-07 | 1.1E-04 | 1.1E-04 |
| Social distancing index | 8.0E-06 | 9.1E-02 | 8.8E-03 | 4.0E-02 | 3.4E-04 | 4.6E-04 | 5.9E-04 | 1.5E-05 | 3.5E-03 | 2.7E-04 |
| Unemployment claims | 4.3E-05 | 6.8E-02 | 1.7E-02 | 1.1E-03 | 2.3E-03 | 8.0E-04 | 1.1E-03 | 2.2E-07 | 4.2E-03 | 3.9E-04 |
| Unemployment rate | 3.7E-08 | 2.6E-02 | 4.4E-03 | 1.7E-01 | 4.2E-04 | 5.3E-06 | 2.4E-04 | 2.8E-03 | 1.5E-04 | 4.8E-05 |
| % working from home | 5.2E-07 | 3.7E-02 | 2.4E-03 | 3.4E-02 | 2.4E-04 | 6.6E-06 | 2.2E-05 | 2.8E-04 | 1.2E-05 | 2.0E-06 |
| Active cases/1000 people | 2.3E-18 | 7.2E-02 | 3.1E-06 | 5.8E-01 | 2.8E-10 | 2.3E-05 | 7.1E-07 | 9.5E-04 | 7.5E-14 | 3.4E-12 |
| Testing capacity | 9.0E-05 | 7.2E-02 | 1.3E-02 | 3.0E-02 | 2.4E-03 | 2.1E-05 | 4.1E-04 | 1.2E-04 | 1.7E-03 | 1.8E-04 |
| Tests done/1000 people | 1.2E-14 | 3.6E-02 | 1.1E-05 | 1.0E-01 | 2.5E-09 | 1.5E-03 | 7.0E-05 | 3.8E-03 | 1.3E-07 | 3.7E-11 |
| Imported COVID cases | 1.3E-16 | 4.5E-01 | 1.1E-04 | 2.6E-01 | 1.7E-07 | 1.3E-03 | 5.1E-04 | 5.9E-04 | 1.7E-06 | 4.4E-12 |
| Retail | 1.3E-05 | 9.6E-02 | 2.2E-02 | 2.9E-02 | 4.4E-03 | 5.9E-04 | 1.4E-03 | 3.4E-07 | 4.5E-03 | 6.1E-04 |
| Grocery | 1.5E-05 | 1.1E-01 | 4.1E-03 | 2.0E-01 | 5.8E-04 | 1.5E-05 | 2.7E-04 | 9.0E-06 | 2.6E-04 | 4.5E-05 |
| Parks | 2.4E-08 | 8.2E-02 | 6.3E-03 | 4.1E-02 | 3.1E-02 | 6.4E-02 | 5.6E-03 | 4.0E-03 | 8.6E-03 | 1.7E-05 |
| Transit | 5.8E-06 | 6.4E-02 | 7.3E-03 | 2.0E-02 | 3.3E-04 | 2.2E-07 | 3.0E-04 | 9.2E-06 | 1.2E-03 | 3.3E-04 |
| Workplaces | 1.5E-05 | 9.6E-03 | 2.6E-03 | 3.5E-03 | 2.1E-05 | 5.8E-08 | 9.3E-06 | 2.2E-05 | 1.2E-04 | 6.1E-05 |
| Residential | 7.0E-05 | 1.7E-02 | 7.2E-03 | 9.6E-05 | 3.6E-04 | 1.7E-07 | 6.5E-05 | 1.1E-06 | 2.8E-04 | 1.5E-04 |
| Protest | 3.6E-01 | 9.6E-01 | 6.6E-01 | 7.7E-01 | 8.0E-03 | 5.1E-01 | 3.8E-01 | 1.7E-01 | 4.1E-01 | 2.3E-03 |
| State | SD | ND | WV | NH | HI | MT | AK | ME | WY | VT |
| Number of cumulative cases | 11559 | 10229 | 9395 | 7150 | 6984 | 6624 | 5666 | 4368 | 3634 | 1572 |
| Miles/person | 3.7E-08 | 1.1E-04 | 1.3E-09 | 3.8E-14 | 9.5E-03 | 5.8E-04 | 2.8E-05 | 8.2E-11 | 2.7E-13 | 4.8E-14 |
| Population | 4.2E-08 | 5.4E-04 | 6.6E-08 | 1.7E-18 | 2.0E-02 | 1.2E-03 | 5.4E-04 | 1.7E-12 | 1.7E-11 | 6.8E-16 |
| % change in consumption | 2.1E-08 | 1.1E-04 | 4.6E-10 | 3.6E-19 | 3.1E-02 | 1.4E-04 | 7.2E-05 | 1.2E-11 | 4.2E-10 | 7.5E-19 |
| Social distancing index | 5.7E-05 | 9.8E-03 | 5.0E-06 | 2.8E-19 | 3.3E-02 | 6.5E-03 | 2.4E-03 | 1.6E-08 | 1.9E-09 | 1.6E-19 |
| Unemployment claims | 5.0E-06 | 4.3E-03 | 1.8E-06 | 3.6E-16 | 3.1E-02 | 3.3E-03 | 1.4E-03 | 1.9E-07 | 1.0E-08 | 2.1E-32 |
| Unemployment rate | 3.8E-08 | 1.3E-03 | 2.5E-06 | 4.8E-26 | 2.9E-02 | 2.7E-03 | 2.8E-04 | 7.1E-18 | 8.0E-12 | 3.8E-14 |
| % working from home | 1.5E-09 | 1.3E-05 | 1.5E-12 | 1.4E-16 | 2.3E-03 | 1.1E-04 | 8.3E-04 | 1.3E-11 | 2.4E-15 | 1.8E-16 |
| Active cases/1000 people | 3.9E-09 | 3.3E-13 | 1.5E-16 | 6.2E-27 | 1.3E-08 | 4.8E-09 | 3.2E-07 | 6.8E-13 | 2.1E-21 | 6.9E-15 |
| Testing capacity | 1.1E-07 | 3.4E-05 | 5.4E-07 | 1.3E-22 | 2.1E-02 | 3.6E-03 | 2.0E-03 | 1.7E-14 | 6.0E-10 | 1.5E-23 |
| Tests done/1000 people | 4.5E-07 | 4.6E-11 | 2.1E-16 | 1.9E-12 | 3.8E-05 | 1.9E-11 | 6.3E-09 | 7.4E-07 | 1.5E-19 | 5.7E-13 |
| Imported COVID cases | 2.7E-07 | 6.8E-09 | 9.6E-13 | 9.0E-32 | 7.4E-06 | 2.7E-08 | 2.0E-06 | 1.3E-14 | 5.4E-19 | 2.5E-13 |
| Retail | 1.3E-07 | 6.1E-03 | 4.4E-06 | 4.8E-19 | 2.9E-02 | 2.7E-04 | 3.1E-04 | 2.0E-09 | 5.4E-14 | 1.4E-20 |
| Grocery | 1.3E-04 | 3.7E-04 | 1.1E-07 | 1.7E-22 | 2.0E-02 | 2.3E-04 | 7.5E-04 | 4.3E-10 | 3.3E-13 | 5.6E-20 |
| Parks | 3.9E-06 | 4.9E-04 | 1.6E-06 | 3.9E-12 | 1.9E-02 | 1.6E-07 | 2.8E-04 | 7.8E-07 | 1.6E-19 | 3.1E-12 |
| Transit | 3.3E-06 | 3.2E-03 | 7.4E-09 | 1.3E-22 | 2.2E-02 | 1.6E-04 | 5.6E-04 | 6.3E-11 | 2.0E-13 | 3.0E-19 |
| Workplaces | 3.9E-12 | 2.3E-04 | 1.2E-07 | 1.4E-23 | 1.8E-02 | 4.6E-04 | 4.7E-04 | 9.8E-15 | 1.9E-09 | 2.2E-19 |
| Residential | 3.5E-09 | 1.6E-03 | 2.0E-06 | 3.0E-24 | 1.6E-02 | 1.4E-03 | 6.3E-04 | 4.9E-12 | 5.6E-08 | 3.1E-22 |
| Protest | 2.4E-01 | 2.6E-01 | 9.7E-01 | 1.1E-03 | 1.3E-02 | 2.1E-01 | 9.6E-03 | 8.0E-02 | 8.3E-01 | 1.6E-01 |

Retail: Retail & recreation, mobility trends for places like restaurants, cafes, shopping centers, theme parks, museums, libraries, and movie theaters.
Grocery: Mobility trends for places like grocery markets, food warehouses, farmers markets, specialty food shops, drug stores, and pharmacies.
Parks: Mobility trends for places like local parks, national parks, public beaches, marinas, dog parks, plazas, and public gardens.
Transit: Transit stations, mobility trends for places like public transport hubs such as subway, bus, and train stations.
Workplaces: Mobility trends for places of work.
Residential: Mobility trends for places of residence.
Test Rate: Ratio of the number of individuals who have taken the virus test over the total population in the region.
Attendee: Number of attendees in the protest.

All 19 metrics showed no significance in causing reduction of the new cases of COVID-19 in Florida. The majority of the 19 metrics did not demonstrate evidence that they can significantly mitigate the spread of COVID-19 in most the affected states such as TX, NY, GA, IL, AZ, NJ, NC, and TN. These 10 states were in the top largest states by population in the US. Public health intervention measures such as closing schools and businesses, avoiding public gatherings, restricting traffic, placing residents to stay-at-home and adherence to guidelines were less well implemented or difficult to implement homogeneously due to large populations and geographical areas (Althouse et al. 2020). These results also explained why the number of new cases of COVID-19 in these states was high and confirmed by several studies (Lai et al. 2020; Masrur et al. 2020; Goldschmidt-Clermont 2020).

Table 3 summarized the ranges of P-values and Table S2 summarized all P-values for testing 18 temporal potential causes of the number of new deaths from COVID-19 across 50 states in the US. All 19 metrics except for protest attendance showed high significant evidence for causing a reduction of new deaths across 50 states except for Michigan (MI) in the US. Our results suggested that a cascade of causes led to the COVID-19 tragedy in the US.

**Table 3.** Ranges of P-values for testing 18 temporal potential causes of the number of new deaths from COVID-19 across 50 states in the US.

| State | P-value |  | State | P-value |  |
| --- | --- | --- | --- | --- | --- |
|  | Lower Bound | Upper Bound |  | Lower Bound | Upper Bound |
| AK | 1.47E-18 | 3.95E-24 | MT | 7.66E-15 | 4.41E-25 |
| AL | 7.45E-11 | 1.10E-18 | NC | 2.93E-08 | 8.95E-18 |
| AR | 1.17E-12 | 1.54E-30 | ND | 2.54E-17 | 1.04E-20 |
| AZ | 4.35E-11 | 1.29E-27 | NE | 5.11E-15 | 2.62E-23 |
| CA | 1.10E-04 | 1.54E-13 | NH | 6.67E-15 | 3.41E-28 |
| CO | 1.41E-21 | 6.44E-26 | NJ | 1.80E-03 | 1.96E-09 |
| CT | 6.73E-07 | 3.13E-16 | NM | 7.20E-07 | 1.15E-16 |
| DE | 1.20E-03 | 1.35E-06 | NV | 9.00E-09 | 1.45E-17 |
| FL | 2.50E-04 | 1.47E-15 | NY | 4.30E-02 | 1.52E-05 |
| GA | 6.53E-07 | 5.81E-17 | OH | 1.82E-08 | 2.67E-17 |
| HI | 1.77E-15 | 1.14E-20 | OK | 4.29E-07 | 1.89E-19 |
| IA | 6.78E-06 | 8.06E-15 | OR | 3.37E-15 | 4.50E-26 |
| ID | 1.96E-08 | 7.11E-18 | PA | 3.86E-15 | 1.11E-23 |
| IL | 3.81E-05 | 1.34E-12 | RI | 4.26E-09 | 2.16E-22 |
| IN | 1.78E-07 | 3.39E-17 | SC | 8.97E-08 | 6.75E-25 |
| KS | 8.74E-30 | 1.42E-40 | SD | 5.04E-18 | 6.01E-25 |
| KY | 1.89E-13 | 1.40E-24 | TN | 7.28E-09 | 2.06E-24 |
| LA | 7.71E-11 | 2.38E-20 | TX | 6.22E-07 | 2.59E-23 |
| MA | 1.25E-03 | 3.92E-04 | UT | 3.90E-13 | 8.12E-29 |
| MD | 3.80E-02 | 7.63E-06 | VA | 3.39E-17 | 2.61E-16 |
| ME | 2.29E-15 | 4.29E-25 | VT | 3.39E-07 | 1.23E-29 |
| MI | 0.28 | 2.07E-05 | WA | 5.67E-16 | 3.83E-29 |
| MN | 2.40E-04 | 8.89E-11 | WI | 2.21E-10 | 4.77E-29 |
| MO | 1.73E-11 | 7.85E-20 | WV | 9.61E-15 | 7.81E-22 |
| MS | 2.94E-08 | 1.49E-19 | WY | 7.93E-26 | 2.39E-28 |

Table 4 listed the most significant risk factor for the new cases of COVID-19 in each of the 50 states in the US. Active Cases/1000 People, workplaces, number of tests completed/1000 people, imported COVD cases, unemployment rate and unemployment claims/1000 people, mobility trends for places of residence (residential), retail & recreation, mobility trends for places like restaurants, cafes, shopping centers, theme parks, museums, libraries, and movie theaters (retail) and test capacity were the most significant risk factors for the new cases of COVID-19 in 23, 7, 6, 5, 4, 2, 1 and 1 states of the US, respectively.

**Table 4.** The most significant risk factor for the new cases of COVID-19 in each of the 50 states in the US.

| P-value | State | Risk Factor | P-value |
| --- | --- | --- | --- |
| 6.27E-09 | MT | Tests Done /1000 People | 1.87E-11 |
| 5.40E-08 | NC | Active Cases/1000 People | 7.49E-14 |
| 4.49E-14 | ND | Active Cases/1000 People | 3.35E-13 |
| 7.12E-07 | NE | Unemployment Rate | 6.10E-10 |
| 2.29E-18 | NH | Imported COVID Cases | 9.04E-32 |
| 1.48E-17 | NJ | Unemployment Claims/1000 People | 2.15E-07 |
| 5.46E-25 | NM | Active Cases/1000 People | 5.68E-13 |
| 3.64E-15 | NV | Imported COVD Cases | 2.36E-09 |
| 9.60E-03 | NY | Residential | 9.57E-05 |
| 2.76E-10 | OH | Active Cases/1000 People | 6.42E-10 |
| 1.31E-08 | OK | Tests Done/1000 People | 4.14E-10 |
| 5.07E-14 | OR | Active Cases /1000 People | 3.87E-12 |
| 4.53E-10 | PA | Work Places | 4.21E-09 |
| 5.82E-08 | RI | Imported COVD Cases | 1.02E-27 |
| 1.12E-09 | SC | Active Cases / 1000 People | 1.58E-04 |
| 1.21E-32 | SD | Work Places | 3.89E-12 |
| 4.30E-26 | TN | Active Cases/1000 People | 3.45E-12 |
| 1.58E-19 | TX | Active Cases/1000 People | 3.05E-06 |
| 3.20E-10 | UT | Active Cases/1000 People | 2.04E-07 |
| 6.35W-09 | VA | Active Cases/ 000 People | 2.08E-08 |
| 7.15E-18 | VT | Unemployment Claims 1000 People | 2.07E-32 |
| 1.16E-10 | WA | Active Cases/1000 People | 2.65E-21 |
| 9.01E-07 | WI | Active Cases/1000 People | 5.19E-12 |
| 2.42E-09 | WV | Active Cases/1000 People | 1.53E-16 |
| 2.16E-12 | WY | Active Cases/1000 People | 2.13E-21 |

Table 5 summarized the most significant risk factor for the deaths from COVID-19 in each of the 50 states in the US. Active Cases/1000 people, workplaces, residential, unemployment rate, imported COVID cases, unemployment claims/1000 people, transit, test done/1000 people, grocery, testing capacity, retail, percentage of change in consumption, percentage of working from home were the most significant risk factor for the deaths of COVID-19 in 17, 10, 4, 4, 3, 2, 2, 2, 1, 1, 1, 1 states, respectively. We also observed that the number of protest attendees showed mild significant evidence to cause increasing the number of new cases of COVID-19 in KY (P-value < 0.00012), KS (P-value < 0.00026), NH (P-value < 0.00108), MA (P-value < 0.0016) and TN (P-value < 0.0024) or to cause more deaths from COVID-19 in OR (P-value < 5.11 E-05), TX (P-value < 0.00017), ME (P-value < 0.00028), KS (P-value < 0.00061), MI (P-value < 0.0015), OH (P-value < 0.0021) and NC (P-value < 0.0023).

**Table 5.** The most significant risk factor for deaths from COVID-19 in each of 50 states in the US.

| Risk Factor | P-value | State | Risk Factor | P-value |
| --- | --- | --- | --- | --- |
| Active Cases/1000 People | 3.95E-24 | MT | Active Case /1000 People | 4.41E-25 |
| Active Cases/1000 People | 1.10E-18 | NC | Work Places | 8.95E-18 |
| Tests Done/1000 People | 1.54E-30 | ND | Unemployment Rate | 1.04E-20 |
| Active Cases/1000 People | 1.29E-27 | NE | Work Places | 2.62E-23 |
| Work Places | 1.54E-13 | NH | Imported COVID Cases | 3.41E-28 |
| Residential | 6.44E-26 | NJ | Residential | 1.96E-09 |
| Active Cases/1000 People | 3.12E-16 | NM | Unemployment Rate | 1.15E-16 |
| Retail | 1.35E-06 | NV | Active Cases/1000 People | 1.45E-17 |
| Active Cases/1000 People | 1.47E-15 | NY | % Change in Consumption | 1.52E-05 |
| Work Places | 5.81E-17 | OH | Unemployment Rate | 2.67E-17 |
| Active Cases/1000 People | 1.14E-20 | OK | Work Places | 1.89E-19 |
| Unemployment Rate | 8.06E-15 | OR | % Working from Home | 4.50E-26 |
| Active Cases/1000 People | 7.11E-18 | PA | Imported COVID cases | 1.11E-23 |
| Residential | 1.34E-12 | RI | Imported COVID cases | 2.16E-22 |
| Work Places | 3.39E-17 | SC | Active Cases/1000 People | 6.75E-25 |
| Grocery | 1.42E-40 | SD | Active Cases/1000 People | 6.01E-24 |
| Work Places | 1.40E-24 | TN | Test Done/1000 People | 2.06E-24 |
| Transit | 2.38E-20 | TX | Active Cases/1000 People | 2.59E-23 |
| Active Cases /1000 People | 3.92E-09 | UT | Active Cases/1000 People | 8.12E-29 |
| Grocery | 7.60E-06 | VA | Work Places | 2.61E-16 |
| Residential | 4.29E-25 | VT | Unemployment Claims/1000 People | 1.23E-29 |
| Unemployment Claims / 1000 People | 2.07E-05 | WA | Transit | 3.83E-29 |
| Testing Capacity | 8.89E-11 | WI | Work Places | 4.77E-29 |
| Work Places | 7.85E-20 | WV | Active Cases/1000 People | 7.81E-22 |
| Active Cases/1000 People | 1.49E-19 | WY | Active Cases/1000 People | 2.39E-28 |

To illustrate the causal relationships between the risk factors and the number of new cases and deaths from COVID-19, we plotted Figures 1 and 2. Figure 1 plotted the social distance index curves as a function of time from March 5, 2020 to August 25, 2020 in Florida (FL) and Rhode Island (RI). Figure 1 showed that the social distance index in FL was much higher than that in RI state, which resulted in the larger number of new cases of COVID-19 in FL than that in RI. Figure 2 showed the number of imported COVID-19 cases as a function of time from March 5, 2020 to August 25, 2020 in Maryland (MD) and Wyoming (WY). We observed a huge difference in the number of imported COVID-19 cases between MD and RI. The very low number of imported cases of COVID-19 in WY resulted in the very low number of deaths from COVID-19 in WY, while the high number of imported cases in MD state led to the increased deaths from COVID-19 in MD. These results were consistent with the finding in the literature. It was reported that strong interventions would substantially decrease the number of deaths (Davies et al. 2020; Gagnon et al. 2020).

**Figure 1.**
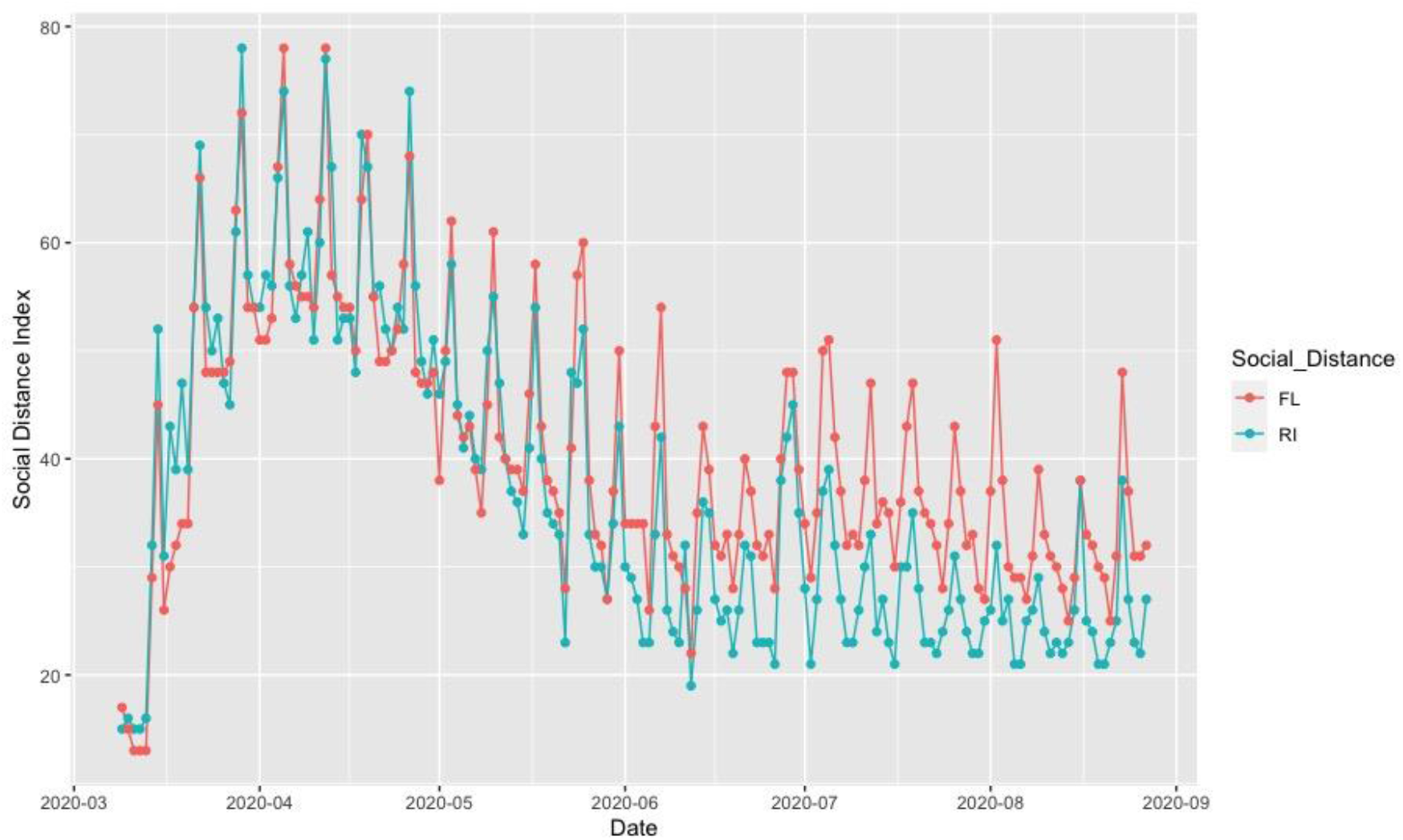
Social distance index curves as a function of time from March 5, 2020 to August 25, 2020 in Florida (FL) (red color) and Rhode Island (RI) (blue color).

**Figure 2.**
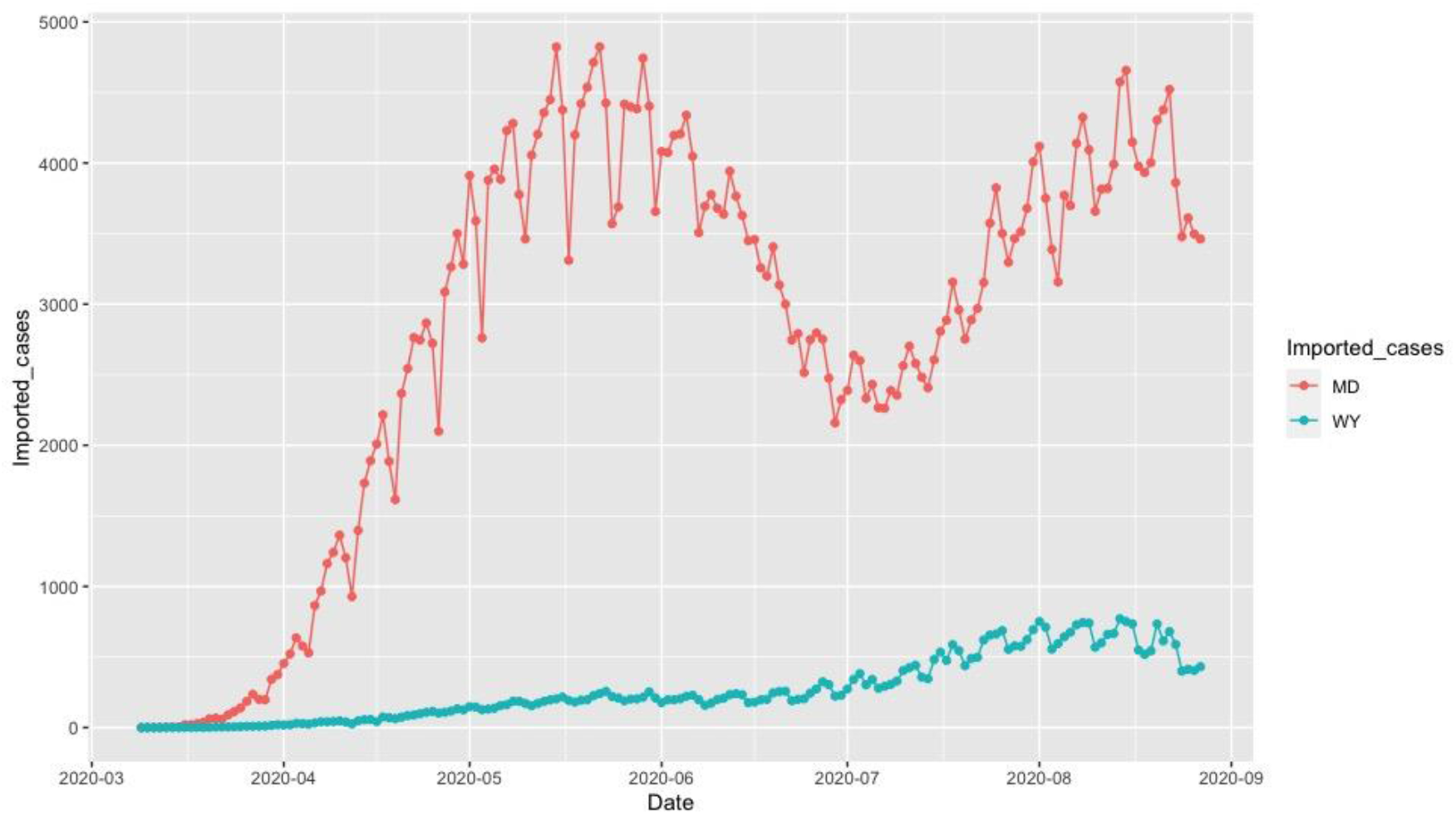
Number of imported COVID-19 cases as a function of time from March 5, 2020 to August 25, 2020 in Maryland (MD) (red color) and Wyoming (WY) (blue color).

## Discussion

Causal inference for COVID-19 is essential for selecting and implementing public intervention measures and understanding the role of the demographics in curbing the spread and reducing the deaths from COVID-19. In this paper, we systematically addressed the issues in identifying causal risk factors and evaluating the causal effects of risk factors and intervention measures on the spread and deaths from COVID-19 in the US. Risk factors and intervention measures included scalar variables and temporal variables. The ANMs were used to test for causal relationships between scalar risk factors and the average number of new cases or deaths from COVID-19 in the US. Transmission of COVID-19 is a dynamic system. Many risk factors and intervention measures are temporal variables. The Granger Causality Test was used to reveal the causal relationships between the temporal risk factors and intervention measures, and the number of new cases or new deaths from COVID-19 across the 50 states in the US.

The demographic risk factors were the major part of the scalar risk factors in the causal analysis of COVID-19. We found that population density was the most significant causal factor of both new cases and death from COVID-19. Population density measured the average number of people per kilometer living in a built-up area. Densely populated states generated conditions where COVID-19 can spread quickly and undetected in the densely populated areas and created high levels of vulnerability. The second significant demographic factor was percentage of males. Our data suggested that men were more vulnerable to Covid-19 than women. However, our analysis did not conclude that more men than women were dying from COVID-19.

We also discovered that more Black Americans were dying from COVID-19. The reasons for this were complex. Black Americans had higher rates of chronic disease conditions, including diabetes, heart disease, and lung disease, were poor and more easily exposed to the COVID-19, and lived in the cramped housing. Inequities in the social determinants of health affected mortality and morbidity of COVID-19 for Hispanic Americans with much milder significance.

We studied the causal effect of major public health interventions across the 50 states in the US. In the absence of centralized intervention measures and implementation of a timeline and presence of the complex dynamics of human mobility and the variable intensity of local outbreaks of COVID-19, evaluating the causal effect of public health intervention measures on COVID-19 transmission and deaths in the USA posed a great challenge. We used 6 Google mobility indexes and 12 daily metrics to measure the effects of COVID-19 spread and public health interventions on mobility and social distancing, derived from mobile device location data and COVID-19 case data, provided by the University of Maryland COVID-19 Impact Analysis Platform. These real time metrics capture the dynamics of social distancing. Granger causality tests were used to identify the causality relationships between time series metrics and time varying in the number of new cases or deaths from COVID-19. Although the risk factors differed by location, Active Cases/1000 people were a significant risk factor for both number of new cases and deaths from COVID-19 in most states. The most popular intervention measure in the US was workplaces (mobility trends for places of work). Workplaces were the significant cause of the number new cases of COVID-19 in 44 states and significant cause of death in 49 states. Therefore, workplaces should be considered as a very important risk mitigation measure to reduce the number of new cases and deaths from COVID-19. Tests done/1000 people was the second population intervention in the US. It was the significant cause of the new cases of COVID-19 in 46 states and significant cause of death in 47 states. Virus test results in quick case identification and isolation to contain COVID-19, and rapid treatment to reduce the number of deaths. Imported COVID cases were also a top significant risk factor for speeding the spread and increasing the deaths from COVID-19. Our results showed that the imported COVID case metric was the significant causal factor for the new cases in 46 states and the significant causal factor for the deaths in 47 states.

Our results showed that the high numbers of cases and deaths from COVID-19 were due to lacking strong interventions and high population density. We observed that no metrics showed significant evidence in mitigating the COVID-19 epidemic in FL and only a few metrics showed evidence in reducing the number of new cases of COVID-19 in AZ, NY and TX. Our results showed strong interventions were needed to contain COVID-19.

Although we tried to systematically and comprehensively analyze the data, this study has multiple limitations. First, we only analyzed the causal relationship between mobility patterns and the number of new cases or deaths and ignored the role of other potential mitigating factors (e.g, wearing face masks) that could also have contributed to the reduction of new cases or deaths from COVID-19. When data are available, more metrics should be included in the analysis.

Second, we have not addressed the confounding bias issue. When confounding is unknown, adjusting for confounding methods cannot be applied to eliminate confounding bias from the causal analysis. Unadjusted confounding bias will distort the inferred (true) causal relationship between the number of new cases or deaths from COVID-19, and metrics for social distancing when these two variables share common causes. This will have substantive implications for developing interventions to mitigate the spread of COVID-19 and reduce the deaths from COVID-19. However, removing confounding from causal analysis for COVID-19 is complicated and will be investigated in the future.

In summary, our analysis has provided information for both individuals and governments to plan future interventions on containing COVID-19 and reduction of deaths from COVID-19.

## Data Availability

All data are publicly available.

https://coronavirus.jhu.edu/MAP.HTML

https://www.google.com/covid19/mobility

https://data.covid.umd.edu

## Acknowledgement

HW Deng was partially supported by NIH grants U19AG05537301 and R01AR069055. Momiao Xiong was partially supported by NIH grants U19AG05537301. The authors thank Sara Barton for editing the manuscript.

